# Evaluation and clinical validation of pan-specific and clade-specific diagnostic real-time PCR assays for monkeypox virus

**DOI:** 10.1101/2025.02.27.25323045

**Authors:** Hong Chang, Tin Hang Hung, Binbin Li, Bobby Lim Ho Kong, Ashwathi Asha Madhaven, Yue Wang, Ming-shan Tsai, Dan Deng, Zhanfeng Cui

**Author notes:** H.C. and T.H.H. contributed equally as co-first authors. Corresponding author: Zhanfeng Cui.

## Abstract

Human mpox, formerly known as monkeypox, has twice been declared a Public Health Emergency of International Concern (PHEIC) by the World Health Organization, in 2022 and 2024. Despite this, global access to rapid, reliable diagnostics remains limited, hindering outbreak control and equitable clinical response. To address this gap, we developed and validated a rapid direct real-time PCR assay capable of detecting and differentiating Clade I and Clade II monkeypox virus (MPXV) directly from clinical samples. We designed a new pan-specific B15L assay to complement the published F3L assay and introduced the first clade-specific assay targeting the B1R gene of Clade I MPXV. In silico validation showed that both B15L and F3L assays detected all available MPXV genomes with 100% sensitivity, while B1R selectively detected only Clade I genomes with 100% specificity. Cross-reactivity screening against 40 high-priority non-target organisms revealed minimal risk of false positives. We confirmed analytical sensitivity of 2 copies per reaction for all assays in vitro, with clade-specific B1R showing 100% specificity. When adapted into a lyophilised direct PCR format for near-point-of-care use, the assay demonstrated detection down to 1 copy per reaction for pan-specific targets and 2 copies for clade-specific detection in spiked clinical samples. This direct PCR assay delivers accurate results in under one hour without requiring nucleic acid extraction. Our combined genomic, laboratory, and clinical validations demonstrate exceptional sensitivity and specificity, positioning this assay as a highly promising tool in emergency mpox diagnostics and a potential model for future outbreak-responsive testing platforms.

## Introduction

Human mpox, formerly known as monkeypox, has emerged as a significant public health concern, particularly highlighted by the World Health Organization’s (WHO) two-time declaration of a Public Health Emergency of International Concern (PHEIC) in May 2022 and August 2024. Infected individuals may experience fever, lymphadenopathy, and a characteristic rash that might lead to secondary infection. Immunocompromised individuals are more vulnerable to severe manifestations of the disease, and in the most severe cases, death. This zoonotic viral disease, caused by the monkeypox virus (*Orthopoxvirus monkeypox*, also known as MPV, MPXV, and hMPXV) first isolated in laboratory monkeys in Denmark in 1958^1^, has recently seen a dramatic increase in global cases, with over 100,000 infections and 230 deaths (accessed 10 November 2024 from https://worldhealthorg.shinyapps.io/mpx_global/^2^).

MPXV has a complex phylogenetic history, primarily categorized into two main clades: Clade I and Clade II. Clade I (formerly Congo Basin or Central African clade), with subclades Ia and Ib, is associated with higher mortality rates, while Clade II (formerly West African clade), with subclades IIa and IIb, has been linked to milder disease outcomes^3–5^. Historically both only led to endemic outbreaks, but the recent global outbreak of mpox since 2022 was caused by an emerging lineage B.1 in the Clade IIb in Europe around May 2022^6,7^, the first PHEIC. This lineage has shown significant genetic divergence and rapid mutation rates, raising concerns regarding its transmissibility and potential for further adaptation^8,9^. Additionally, the emergence of Clade Ib, which causes a more widespread endemic outbreak in the Democratic Republic of Congo, has shown increased human-to-human transmission^10^ and thus the second PHEIC. In November 2024, United Kingdom confirmed the first local transmission of Clade Ib mpox outside Africa since the onset of second PHEIC^11^.

The emergence of new strains is particularly alarming as they may exhibit enhanced transmissibility and virulence, posing a substantial public health threat^12^. WHO has already added MPXV to the list of priority pathogens that might cause a pandemic. The global spread of Clade IIb, coupled with the escape of Clade I from endemic areas, underscores the necessity for vigilant surveillance to mitigate and contain the risk of a pandemic.

Molecular diagnostics plays a crucial role in mitigating the risk of pandemic by enabling mass screening and rapid identification, as we learned from the immediate past lesson from the COVID-19 pandemic^13^. Early evidence, such as from Switzerland, demonstrated that rapid turnaround of molecular diagnostics in combination with effective contact tracing was essential in outbreak control^14^. Among various diagnostic methods available, real-time polymerase chain reaction (RT-PCR) assays remain the gold standard for its high sensitivity and specificity, especially useful for low target level when pathogen loads may still be minimal in early stages of infection^15^. The quantitative nature also offers significance in monitoring viral load and estimating infectivity^16^. However, traditional real-time PCR systems require expensive equipment and highly trained professionals to operate, significantly limiting their use to well-equipped laboratories.

The Target Product Profiles (TPP) for published by WHO^17^ in 2023 has highlighted the pressing need for more affordable, simplified, and decentralized diagnostic kits for mpox in outbreak regions. While several commercial mpox real-time PCR kits have been available since the PHEIC declaration in 2022, all currently require DNA extraction and purification steps to eliminate PCR inhibitors commonly found in clinical samples, only suitable for clinical laboratories when handling large volume of tests^18^. Moreover, none of the kits approved under the WHO Emergency Use Listing (EUL) or CDC Emergency Use Authorization (EUA) can differentiate between MPXV Clades I and II, despite this being one of the preferred characteristics outlined in the WHO mpox TPPs.

The overarching goal of this paper is to develop clade-specific diagnostic primers and probes for integration into real-time PCR assay. First, we describe how we optimise the primer design algorithm based on the openly available genomic data. Second, we evaluate their sensitivity and specificity in differentiating the MPXV clades in both *in silico* and *in vitro* tests. Third, we validate the performance of our assay on human clinical samples that were spiked with synthetic MPXV DNA fragments and evaluate its analytical sensitivity.

## Methods

### MPXV pan-specific primer design and screening

We downloaded all complete genomic data of MPXV (NCBI Taxon: 10244) using NCBI Datasets. We also followed the WHO instructions and requirements for emergency use listing submission on *in vitro* diagnostic testing of MPXV nucleic acid (PQDx_457 version; 10 September 2024) in obtaining a list of 40 non-target high priority organisms (**Supplementary Table 1**), including same-family viruses such as smallpox virus, camelpox virus, and buffalopox, and other common contaminants such as herpes simplex virus, streptococcus, and human genomic DNA.

We mapped all MPXV genomic sequences to the reference genome NC_003310.1 and conducted multiple sequence alignment using ViralMSA 1.1.30. We then designed mpox-specific primer using the real-time PCR mode in varVAMP 1.2.1 mode and screened primers that might bind to non-target genomes using blastn 2.16.0. All candidates for primers and probes were aligned to the reference and non-target genomes to inspect their sequence similarity and predict their binding efficiency. We picked the highest-confidence primers and probes targeting the **B15L** loci and obtained the primers and probes synthesised by Integrated DNA Technologies (United Kingdom). We also included another previously designed primer scheme on **F3L**^19^. Sequences and statistics for all primers and probes listed in this study could be found in **Supplementary Table 2**.

### MPXV clade-specific primer design

With reference to the previous phylogenetic study^20^, 56 MPXV genomes were chosen for the clade-specific primer design. Among the 56 genomes, 2 of them were reference genomes for Clade I (NC_003310) and Clade II (NC_063383) MPXV, while the other 54 were public genomic resources in NCBI (**Supplementary Table 3** of their accession & origin & clades in supp). We aligned the genomes by using MAFFT version 7 (https://mafft.cbrc.jp). Clade-specific regions were screened based on an indel observed in the B1R loci based on the multiple sequence alignment, where an ∼2.5 Kb-insertion in Clade II would prevent real-time PCR amplification. We manually designed clade-specific primers and probes that flanked the **B1R** loci.

### Homology analysis of specificity

We analysed the sequences of primers and probes of B15L, F3L, and B1R for homology with full-length genomic sequences attained from Nextstrain. Nextstrain contained 473 Clade Ia sequences, 128 Clade Ib sequences, 25 Clade IIa sequences, and 221 Clade IIb sequences. We ran blastn 2.16.0 with the following command due to the nature of primer and probe sequences: ‘-task blastn-short -word_size 7 -evalue 100 -perc_identity 80 -reward 2 - penalty -3 -gapopen 2 -gapextend 1 -dust nò, which increased sensitivity for short sequences, retained alignments with weaker statistical significance, and permitted small indels with minimal penalties. We also manually crosschecked the homology results and the sequences and removed those that contained significant number of unknown bases, as stretches of ‘N’, in our target amplicons. The final analyses contained 393 Clade Ia sequences, 55 Clade Ib sequences, 25 Clade IIa sequences, and 217 Clade IIb sequences.

### In *silico* inclusivity test for MPXV pan-specific primers

We used R version 4.4.0 to analyse the homology results and potential amplification successes. First, all the homology hits from blastn were sorted based on the genomic coordinates for each of the two primer sets (B15L and F3L). Then, potential amplification success was determined if the hits from forward primer, probe, and reverse primer, or the reverse way, were consecutive.

We then visualised two statistics. First, we calculated the sensitivity, which was calculated as the number of potential amplification success divided by the total number of genomes tested, for each of the four sub-clades. Second, for those with potential amplification success, we calculated the sequence identity for each genomic sequence on the four sub-clades.

### In *silico* specificity test for MPXV clade-specific primers

We ran the same blastn command as above using B1R Clade I-specific primers and probe with the same full set of MPXV genomic sequences and the same R analysis as above to analyse potential amplification successes.

We determined potential amplification successes by both the consecutive homologies of forward primer, probe, and reverse primer, and the potential amplicon length. While there is no absolute consensus on the limit of amplicon length by real-time PCR, most guidelines recommend < 200 bp for optimal efficiency. We used < 1,000 bp as the criterion for potential success.

### In *silico* cross-reactivity test with other non-target organisms

We analysed the sequences of primers and probes for homology with full-length genomic sequences for 40 non-target high-priority organisms as above. We searched the meta data of complete genomes for each of the organisms using their NCBI Taxonomy IDs with NCBI Datasets command-line tools version 16.29.0. We then randomly selected 5 representative genomes for each organism and merged all the genomes into a single sequence file. List of randomly selected representative genomes could be found in **Supplementary Table 4**. We ran blastn 2.16.0 with the same parameters as above.

We then used R version 4.4.0 to analyse the homology results and potential amplification successes as above.

We then visualised two statistics. First, we calculated the frequency of potential homology, which was calculated as the number of homology hits from blastn divided by the total number of genomes tested, for each of the four sub-clades. Second, we reported the average sequence similarity.

### In *vitro* MPXV direct real-time PCR assay

For MPXV quantification in subsequent *in vitro* analyses, real-time PCR assays were performed with 20-µl reactions, which contained 10 µl 2× Lyo PCR Master Mix (Apto-Gen Ltd, United Kingdom), 5 µl primer-probe pre-mix (200nM B15L and F3L pan-specific primers and 100nM probe; or 400nM B1R Clade I-specific primer and 200nM probe), and 5 µl samples (viral samples or synthetic DNA standards), using a QuantStudio 5 Real-Time PCR System (Thermo Fisher Scientific, USA). The thermocycling profile for RT-PCR was as follows: an initial denaturation at 95°C for 1 min, followed by 40 cycles of 95°C for 10 s and 60°C for 15 s. Signal was collected after each cycle.

### Synthesis of MPXV genomic standards and establishment of standard curve

We designed two synthetic standards (Integrated DNA Technologies, United Kingdom) for MPXV genomes using the multiple sequence alignment obtained from above: an 877-bp Clade I-like sequence that contained sequences of F3L, B15L and B1R genes; and a 618-bp Clade II-like sequence that contained sequences of F3L, B15L and D13L genes. The sequences of the MPXV synthetic standards can be found in **Supplementary Table 5**.

Synthetic MPXV genomic DNA standards were first diluted by 1X IDTE buffer (Integrated DNA Technologies, United Kingdom) to 10 ng/µl. Accurate concentration of the stock DNA standard was then determined by Qubit dsDNA Quantification Broad Range Assay Kit (Thermo Fisher Scientific, USA). DNA copy number and dilution volume were calculated with Thermo Scientific Web Tools. Then, DNA standards were serially diluted (10-fold, from 10^6^ to 10^0^ copies/µl) and real-time PCR was performed on QuantStudio™ 5 Real-Time PCR System (Thermo Fisher Scientific, USA). Experiment was triplicated for each concentration, and the mean and standard deviation of the C_t_ value was calculated. In order to establish the standard curve, the mean of C_t_ was plotted against the log_10_ (DNA copy number), with error bars showing the standard deviation. By using the model of linear regression, the equation, hence the slope (*m*) and goodness of fit (*R^2^*) of the standard curve were obtained. The amplification efficiency (*E)* was calculated by using the following formula: *E* = [10^(−1/*m*)^ – 1] × 100%.

### In *vitro* sensitivity for MPXV primers

We first evaluated the analytical sensitivity of each primer set (pan-specific B15L and F3L and Clade I-specific B1R) by determining the tentative limit of detection (LoD), which was defined as the lowest amount of target compound in a sample that could be accurately measured by the primer. We adopted the CLSI EP17-A2 guideline, which sets the standard for evaluation and documentation of the detection capability of clinical laboratory measurement procedures^21^. Real-time PCR reaction was run for each set of primer pairs. Experiment was triplicated with a range of synthetic DNA concentrations (2 × 10^4^, 2 × 10^3^, 2 × 10^2^, 2 × 10^1^, 2 × 10^0^ and 2 × 10^−1^ copies/reaction). Tentative LoD was estimated when positive detection was below 100%. To confirm the experimental sensitivity of each primer, experiment was repeated for 20 replicates, with sample concentration at the tentative LoD. The analytical LoD was confirmed when MPXV was positively detected in 95% of the replicates.

### In *vitro* specificity of MPXV pan-specific and Clade I-specific primer

In order to determine the experimental specificity of MPXV pan-specific and Clade I-specific primer, real-time PCR assays were performed with 2,000-copies MPXV Clade II synthetic standard. Specificity of MPXV pan-specific and Clade I-specific primer (B1R) was then estimated by the numbers of true positives and false negatives.

### Acquisition and preparation of clinical matrix

We obtained 50 human skin lesion swab samples from the Department of Dermatology, Shanghai Children’s Medical Center, School of Medicine, Shanghai Jiao Tong University. Details of the sample types could be found in **Supplementary Table 6**. This study was approved by the Institutional Review Board of Shanghai Children’s Medical Center (SCMCIRB-K2025018-1). Anonymised clinical samples were provided to Oxford Suzhou Centre for Advanced Research for processing.

To prepare the MPXV-negative clinical matrix, 500 µL of RNase-free water was added to each of the 50 swab tubes, which were then vortexed for 10 minutes. Then, 200 µl of supernatant from each tube was pooled into a single 50-ml centrifuge tube, and the total volume was adjusted to 10 ml. Finally, the matrix was aliquoted and stored at –80°C.

### Clinical validation of analytical sensitivity with spiked standards and irradiated virus

We incorporated our assay into the UlfaQ™ Direct PCR system (ZYTCA Ltd, United Kingdom) (**Figure 1**), which contained an enhanced reaction buffer that eliminated effects of inhibitors. We added 16 µl of MPXV-negative clinical matrix and 25 µl UlfaQ Sample Processing Control (SPC) into 500 µl of UlfaQ lysis buffer, then spiked in 4 µl of our MPXV synthetic standard. We included standards of six different concentrations (150, 125, 100, 75, 50, and 0 copy/ml) and a negative control (without SPC). We tested Clade I and Clade II separately thus in total we had 14 matrices.

**Figure 1.**
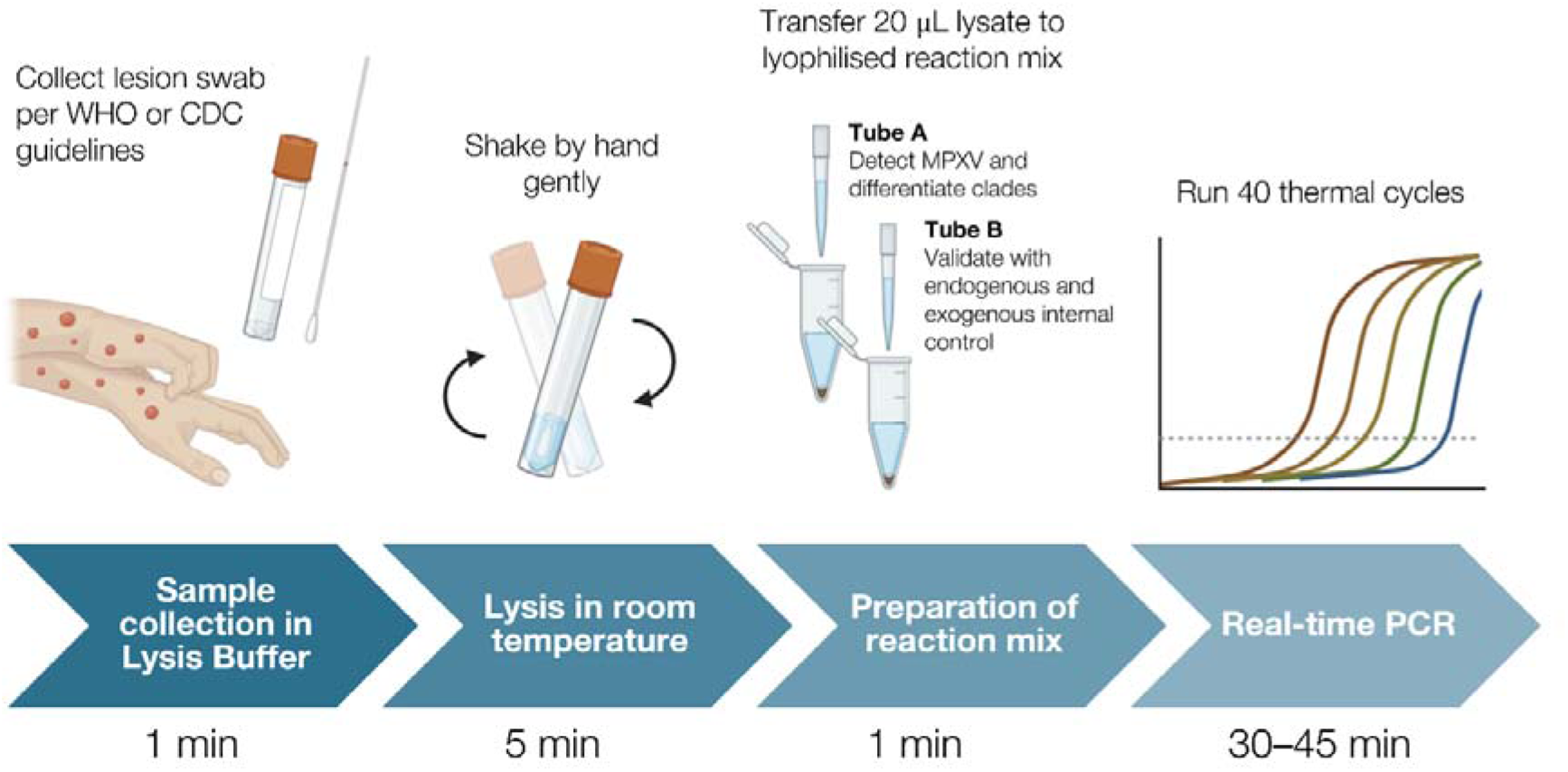
Graphical abstract of direct real-time PCR in clinical validation.

For each matrix, we transferred 20 µl of the lysate into a pre-lyophilised reaction master mix (Tube A) which contained UlfaQ™ Direct PCR mix and our MPXV primers and probes. Another 20 µl of the lysate was also added in the lyophilised internal control assay (Tube B) targeting SPC and human beta-actin gene. We performed RT-PCR on three replicates for each matrix on QuantStudio™ 5 Real-Time PCR System (Thermo Fisher Scientific, USA). The thermocycling profile for real-time PCR was as follows: an initial denaturation at 95°C for 1 min, followed by 40 cycles of 95°C for 5 seconds and 60°C for 10 seconds.

The analytical sensitivity of our assay was determined in similar approach of CLSI EP17-A2 guideline^21^ as described above. Tentative LoD was estimated when positive detection was below 100%. To confirm the analytical sensitivity of each primer, experiment was repeated for 20 replicates, with sample concentration at the tentative LoD. The analytical sensitivity was confirmed when MPXV was positively detected in 95% of the replicates.

As a complementary analysis, LoD_95_ was estimated using probit regression. Binary detection outcomes were modelled against input number of copies per reaction using a generalized linear model with probit link. LoD_95_ was defined as the concentration corresponding to a predicted 95% detection probability.

In an additional experiment, we replaced the MPXV synthetic standard spike-ins with irradiated MPXV Clade II virus (2301231v) obtained from the Culture Collections of UK Health Security Agency, at six different concentrations (1000, 500, 250, 125, 62.5, 31.25 TCID_50_/ml, where TCID_50_ stands for the median tissue culture infectious dose). We ran the same qPCR assay and determined the tentative LoD and analytical sensitivity.

## Results

### In *silico* inclusivity test for MPXV pan-specific primers (B15L and F3L)

We found that both B15L and F3L primers were highly successful in amplifying MPXV across the four sub-clades (**Figure 2a**). Both assays achieved 100% *in silico*-predicted sensitivity for MPXV across all subclades, including 393 Clade Ia, 55 Clade Ib, 25 Clade IIa, and 217 Clade IIb genomes.

**Figure 2.**
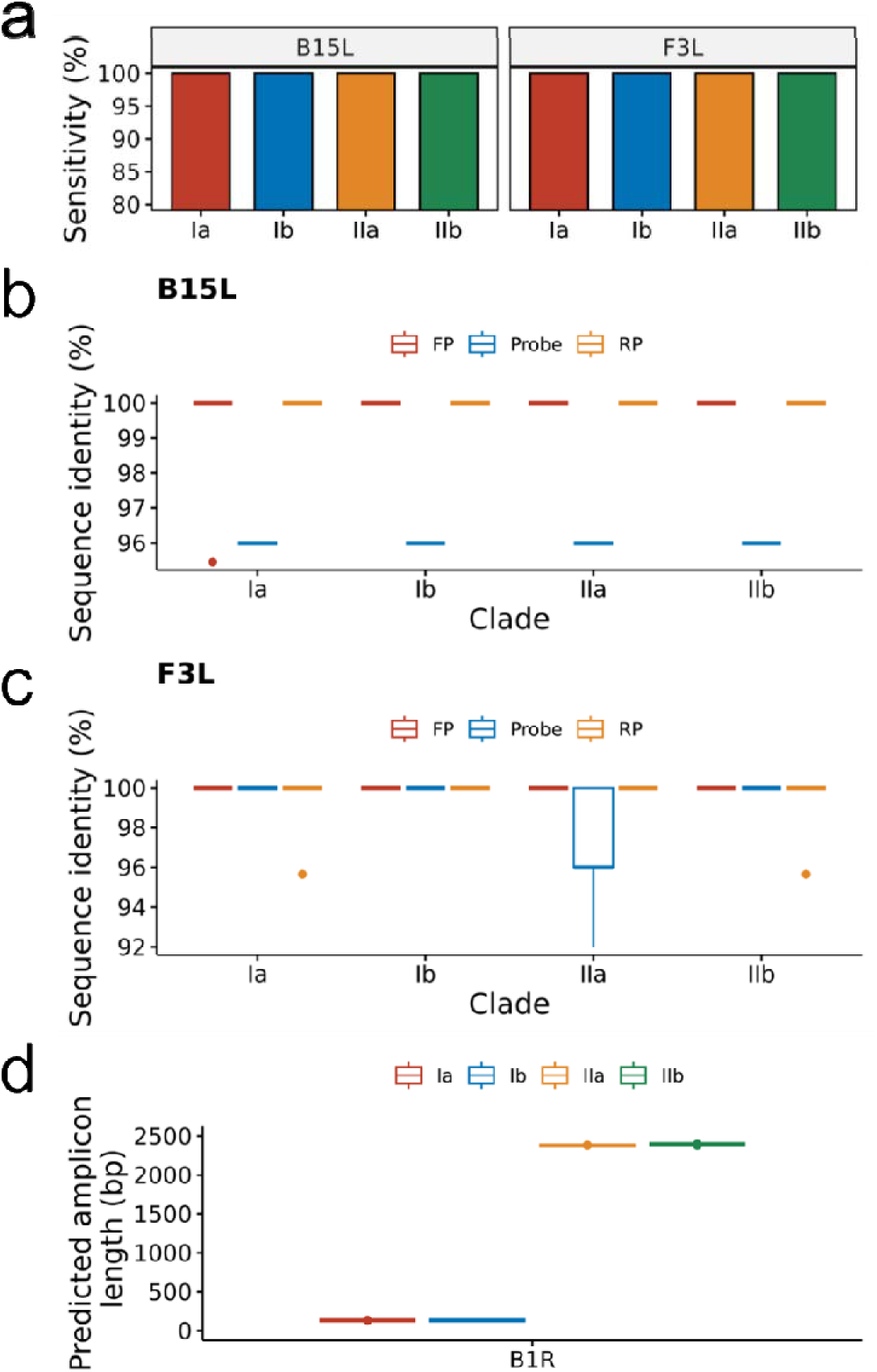
**(a)** *In silico-*predicted sensitivity for B15L and F3L across four MPXV sub-clades, calculated as number of potential amplification successes divided by total number of genomic sequences tested; and sequence identity for forward primer (FP), probe, and reverse primer (RP) for **(b)** B15L and **(c)** F3L across four MPXV sub-clades and two genomic databases (GISAID and Nextstrain). **(d)** Potential amplicon length using B1R primers across four MPXV sub-clades.

We found > 90% sequence similarity for all primers and probes (**Figure 2b and c**). Specifically, all B15L and F3L primers had 100% sequence similarity with the genomic targets, except 3 outliers for each. B15L probe had a consistent 96% sequence similarity with the genomic targets across all sub-clades. F3L probes had a consistent 100% sequence similarity with the genomic targets across all-subclades except the median sequence similarity for sub-Clade IIa was 96%.

### In *silico* sensitivity and specificity test for MPXV clade-specific primers (B1R)

We found that B1R primers and probes were highly specific for MPXV Clade I (**Figure 2d**). The median predicted amplicon length for both sub-Clade Ia was 134 bp and Ib was 134 bp and that for sub-Clade IIa and IIb was 2,379 bp and 2,398 bp respectively. All 393 Clade Ia and 55 Clade Ib were predicted to amplify, while none of the 25 Clade IIa and 217 Clade IIb genomes were predicted to amplify. Thus, the sensitivity and specificity for B1R assay was 100%.

### In *silico* cross-reactivity test with other non-target organisms

We analysed the sequences of primers and probes for homology with full-length genomic sequences for 40 non-target high-priority organisms as above. Homology was found between primers and Poxviridae viruses, more specifically, buffalopox, camelpox, cowpox, and mousepox viruses (**Figure 3a–c**), where some primers were found to have homology hits in blast in all genomes. However, for the vast majority of them the average sequence similarity was sufficiently low, except for B1R forward primer, which shared a 96% similarity with all buffalopox, cowpox virus, and mousepox viruses (**Figure 3d–f**).

**Figure 3.**
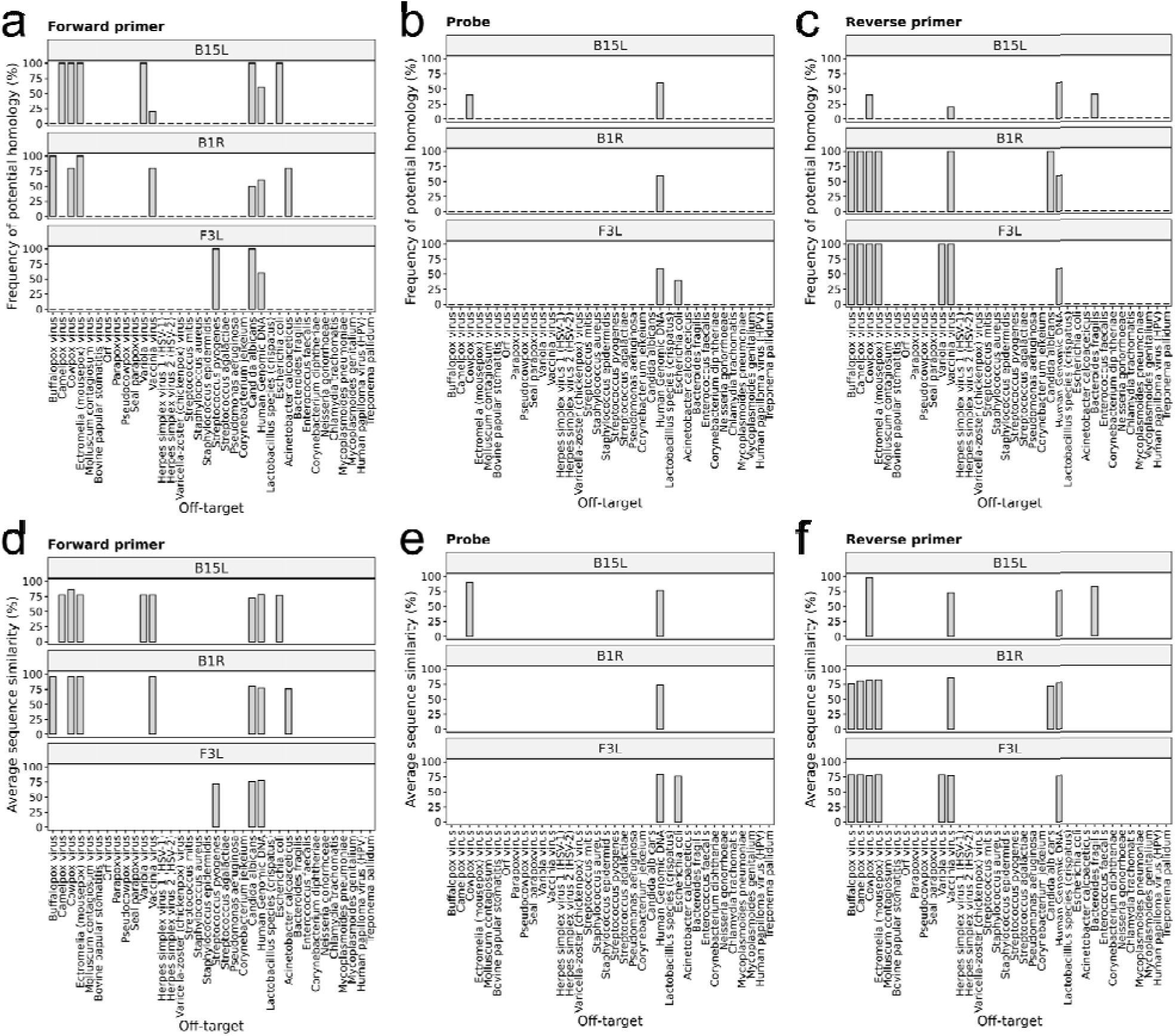
Frequency of homology hits between **(a)** forward primer, **(b)** probe, and **(c)** reverse primer for B15L, B1R, and F3L and the genomic sequences of each non-target organism (*N =* 5, except for organisms that did not have five representative genomes available). List of non-target organisms was determined from the relevant WHO guideline on mpox testing. Average sequence similarity between **(d)** forward primer, **(e)** probe, and **(f)** reverse primer for B15L, B1R, and F3L and the genomic sequences of each non-target organism (*N* = 5, except for organisms that did not have five representative genomes available). List of non-target organisms was determined from the relevant WHO guideline on mpox testing.

We predicted that the only potential off-target amplification would be B15L on cowpox virus. Out of the five genomes tested, two genomes (HQ407377.1 and KY369926.1) showed >80 % sequence similarity with all forward primer, probe, and reverse primer of B15L (**Supplementary Table 7**).

### Standard curve and assay performance

MPXV Clade I synthetic DNA was used to create standard curve and evaluate the performance of the real-time PCR assay. The detection of the real-time PCR reactions was linear over six 10-fold dilutions (2 × 10^6^ to 2 × 10^1^ copies/reaction) with a correlation coefficient 1.0, for both MPXV pan-specific primers and Clade I-specific primer (**Figure 4a** and **Supplementary Table 8**). Due to the high variance of Ct value at low copy number, the data of Ct value at 2 copies/reaction was omitted in plotting the standard curve. Amplification efficiency was high for both primer sets, in which all of them were 90–110%. No false positive was observed in the triplicated reactions, indicating the reliability of the proposed real-time PCR assays.

**Figure 4.**
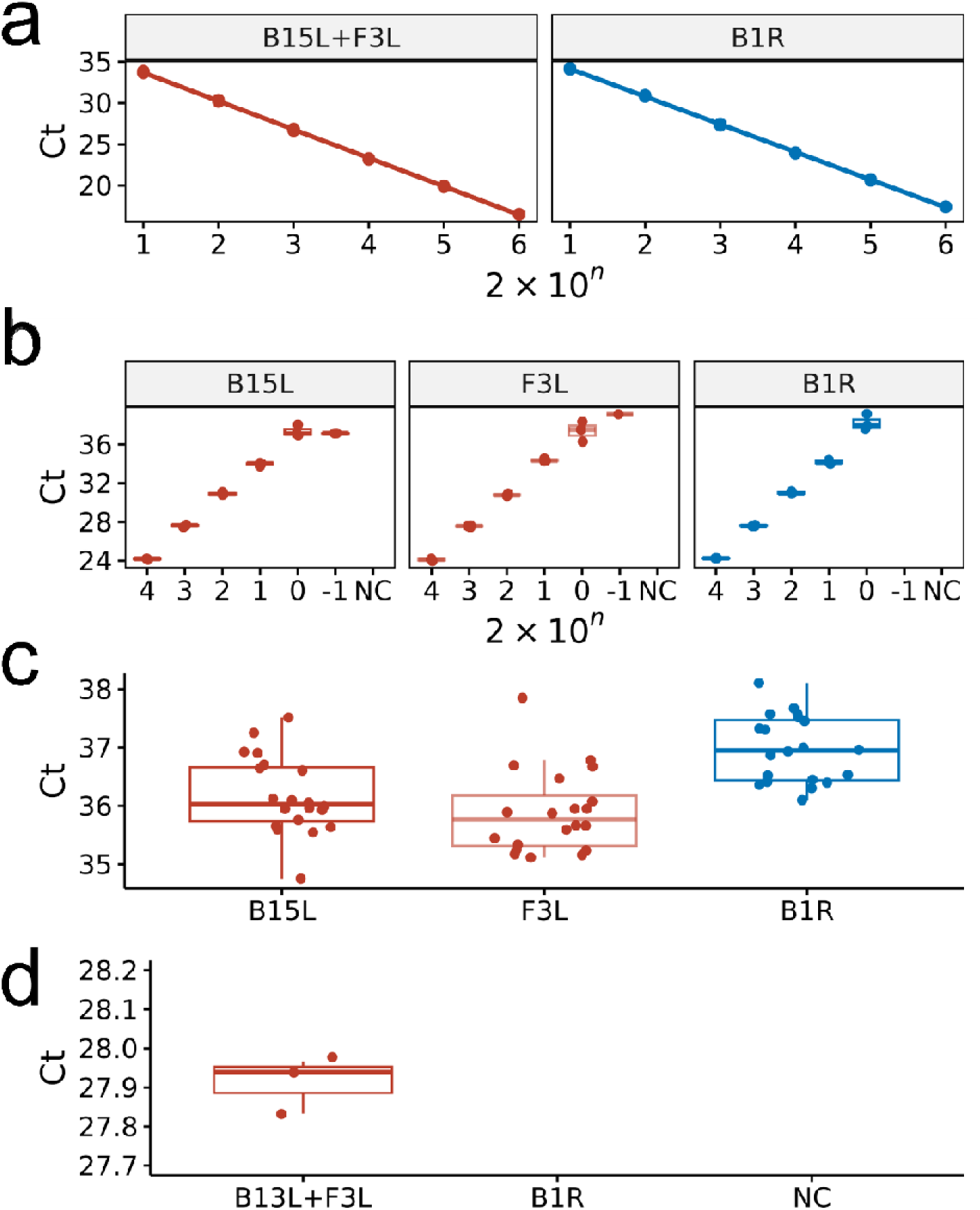
**(a)** Standard curve of real-time PCR Ct value against copy numbers of MPXV synthetic standard for pan-specific B15L+F3L primers and clade-specific B1R primers. Experiment was triplicated. **(b)** Tentative limit of detection (LoD) determined by real-time PCR Ct value against copy number of MPXV synthetic standard. Experiment was triplicated. **(c)** Analytical sensitivity confirmed by real-time PCR of 20 replicates at the tentative LoD. **(d)** Analytical specificity of Clade 1-specific B1R on MPXV Clade II synthetic standard. NC denotes negative control.

### In *vitro* analytical sensitivity for B15L, F3L, and B1R

To determine the tentative LoD of each primer set in standard laboratory settings, real-time PCR assays were triplicated with MPXV synthetic standard over six concentrations (2 × 10^4^, 2 × 10^3^, 2 × 10^2^, 2 × 10^1^, 2 × 10^0^ and 2 × 10^−1^ copies/reaction). All primer sets could successfully amplify the target at an input amount of 2 copies, the tentative LoD (**Figure 4b** and **Supplementary Table 9**). To confirm the experimental sensitivity of each primer set, all primer sets demonstrated 100% sensitivity when repeated for 20 reactions at an input amount of 2 copies (equivalent to 100 copies/ml), confirming the analytical LoD in standard laboratory condition (**Figure 4c** and **Supplementary Table 10**).

### In *vitro* specificity of MPXV Clade I-specific assay B1R

To evaluate the specificity of MPXV Clade I-specific primer, real-time PCR was performed by using MPXV Clade II synthetic standard. Pan-specific assay (B15L+F3L) successfully detected 2,000 copies of MPXV Clade II synthetic standard, while Clade I-specific primer (B1R) showed no amplification (**Figure 4d**). The triplicated experiment confirmed the specificity of MPXV Clade-I specific primer to be 100%.

### Clinical validation of analytical sensitivity

For MPXV Clade I synthetic standard, we successfully detected positive signals in all triplicates at 3, 2.5, 2, 1.5, and 1 copies/reaction for B15L+F3L pan-specific assay and at 3, 2.5, and 2 copies/reaction for B1R Clade I-specific assay (**Figure 5a** and **Supplementary Table 11**). We thus estimated the tentative LoD for Clade I to be 2 copies/reaction (equivalent to 100 copies/ml). We further validated the LoD with 20 replicates. All replicates gave positive signals and thus confirmed the analytical sensitivity with high confidence (**Figure 5b** and **Supplementary Table 12**).

**Figure 5.**
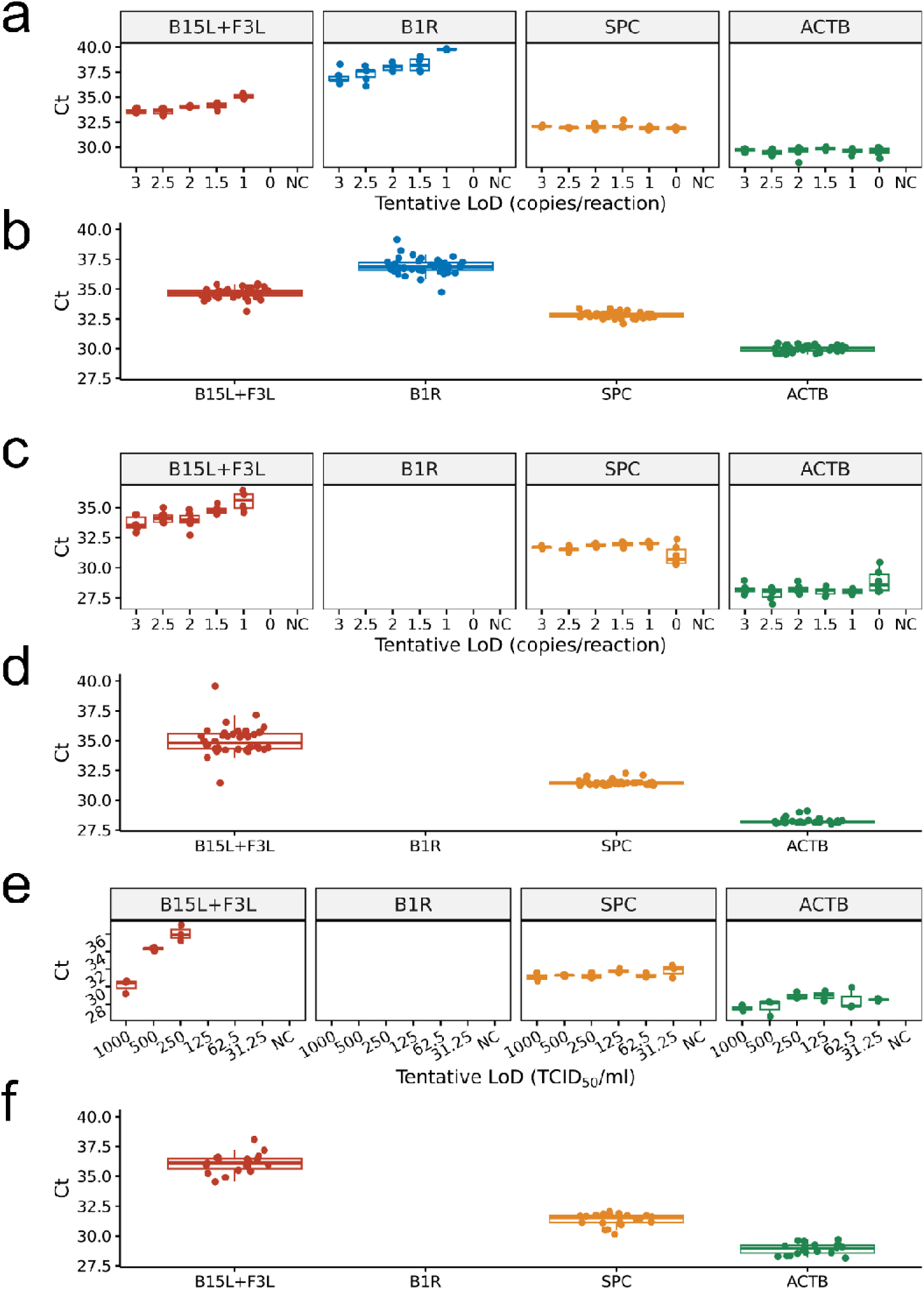
**(a)** Tentative limit of detection (LoD) for MPXV Clade I synthetic standard determined by real-time PCR Ct value against copy number of standard. Experiment was triplicated. **(b)** Analytical sensitivity for MPXV Clade I synthetic standard confirmed by real-time PCR of 20 replicates at the tentative LoD. **(c)** Tentative limit of detection (LoD) for MPXV Clade II synthetic standard determined by real-time PCR Ct value against copy number of standard. Experiment was triplicated. **(d)** Analytical sensitivity for MPXV Clade II synthetic standard confirmed by real-time PCR of 20 replicates at the tentative LoD. **(e)** Tentative limit of detection (LoD) for MPXV Clade II irradiated virus determined by real-time PCR Ct value against TCID_50_ (median tissue culture infectious dose). Experiment was triplicated. **(f)** Analytical sensitivity for MPXV Clade II irradiated virus confirmed by real-time PCR of 20 replicates at the tentative LoD.

For MPXV Clade II synthetic standard, we successfully detected positive signals in all triplicates at 3, 2.5, 2, 1.5, and 1 copies/reaction for B15L+F3L pan-specific assay (**Figure 5c** and **Supplementary Table 13**). We thus estimated the tentative LoD for Clade II to be 1 copy/reaction (equivalent to 50 copies/ml). We further validated the LoD with 20 replicates. All replicates gave positive signals and thus confirmed the analytical LoD with high confidence (**Figure 5d** and **Supplementary Table 14)**.

Probit regression models estimated even lower LoD. For MPXV Clade I, the regression-estimated LoD_95_ was 0.63 copies/reaction for the pan-specific B15L+F3L assay and 1.88 copies/reaction for the clade-specific B1R assay. For MPXV Clade II, the regression-estimated LoD_95_ for the B15L+F3L assay was 0.63 copies/reaction. However notably, for the pan-specific assay, detection transitioned sharply from 0% at 0 copies/reaction to 100% at ≥1 copy/reaction. Under such near step-function behaviour, regression-based LoD values are mathematically interpolated between discrete experimental concentrations and therefore reflect model-derived estimates rather than directly observed detection thresholds. It was infeasible and biologically irrelevant to have standards below 1 copy/reaction to better estimate regression-based LoD, therefore the primary LoD values reported in this study follow EP17-A2 verification.

For MPXV Clade II irradiated virus, we successfully detected positive signals in all triplicates at 1000, 500, and 250 TCID_50_/ml for B15L+F3L pan-specific assay (**Figure 5e and Supplementary Table 15**). We thus estimated the tentative LoD for Clade II to be 250 TCID_50_/ml. We further validated the LoD with 20 replicates. All replicates gave positive signals and thus confirmed the analytical LoD with high confidence (**Figure 5f and Supplementary Table 16**).

None of the replicates at any concentration of Clade II standard gave positive signals for B1R Clade I-specific assay, thus confirming the perfect specificity to differentiate MPXV clades.

## Discussion

This study presents an integrated pan-specific and clade-specific diagnostic assay suitable for near-point-of-care detection of human mpox. The assay has been validated through in silico analyses using a comprehensive genomic database, and through in vitro experiments using synthetic standards, including a test of analytical limit of detection (LoD).

Our assay included two new target regions, pan-specific B15L and clade-specific B1R, and a verified pan-specific F3L. The function of B15L (IL-1 beta inhibitor) gene is not yet reported but other IL-1 beta inhibitors might interfere with host cell signalling pathways, further aiding the virus in evading immune detection and response^22^. The B1R gene encodes for a BTB-Kelch-domain protein and has a frameshift mutation and a within-gene deletion in Clade I MPXV^23^, and thus initiates translation at a different start codon between Clade I and IIb MPXV, resulting in different fragments of the protein. It is also suggested a close homologue of B1R gene might inhibit the NFκB pathway and stimulates CD8+ T cell proliferation but such a function in MPXV is yet to be confirmed^24^. The F3L encodes an enzyme with a double-stranded RNA binding domain and might be responsible for a broad spectrum of antiviral immune responses^25^ and circumventing apoptosis to achieve productive replication^24^ in the host.

Through *in silico* approaches, we have validated that the MPXV pan-specific assays targeting B15L and F3L showed perfect sensitivity to historical and contemporary MPXV genomes across different lineages of sub-clades. Most importantly, our clade-specific B1R assay is predicted to have high sensitivity and specificity to Clade I MPXV. Therefore, when combined with use of either B15L and F3L as the pan-specific assay, our approach allows for clade differentiations, where Clade I would yield positive signals for both B1R and B15L/F3L, and Clade II would be B1R-negative but B15L/F3L-positive. The diagnostic assay was made most efficient in a duplex real-time PCR, where each probe can be labelled with a different fluorophore (i.e. FAM and HEX). Although not tested in real-life outbreak settings, our direct real-time PCR, clade-specific assay would be directly addressing a significant gap in disease surveillance, as there is still no WHO-approved antigen-based RDT or point-of-care tests that can distinguish between mpox clades^26^. The rapid turnaround time (<1 hour) offers potential to reduce time and financial cost for clade differentiation in central laboratories.

We also thoroughly ruled out potential cross reactivity with other closely related viruses and common contaminants in clinical samples. All B15L, F3L, and B1R showed a very high degree of specificity even among evolutionary relatives, such as buffalopox, camelpox, cowpox, and mousepox viruses. Occasional in silico matches to B15L sequences in cowpox genomes were observed. However, cowpox is very rare and only a few hundred cases have ever been reported^27^. There is also no reported case of human-to-human transmission^28^. Therefore, a false positive result due to cowpox infection would be highly unlikely. Nonetheless, in clinical settings where orthopoxviruses may co-circulate, confirmatory testing should be considered in the case of positive results. We thus remain confident that our B15L, F3L, and B1R assays demonstrate excellent sensitivity and specificity to MPXV.

The analytical sensitivity of our assay, 2 copies per reaction for MPXV Clade I and 1 copy per reaction for MPXV Clade II, demonstrated utility in detecting extremely low amount of MPXV genomes. Our sensitivity is more superior than any assay reported on mpox, including Li et al.’s at 3.5–40.4 copies^29^, Maksyutov et al.’s at 20 copies^19^, Li et al.’s at 15 copies^30^, Mills et al.’s at 3.3 copies^31^, and Paniz-Mondolfi et al.’s at 7.2 copies^32^, except Uhteg and Mostafa’s also at 2 copies but theirs could not differentiate clades^33^. It should be noted, however, that these LoD values were derived under controlled experimental conditions using synthetic material, and actual performance may vary in complex clinical matrices. We also further validated the sensitivity and specificity of our assay using irradiated virus at 250 TCID_50_/ml. The primary LoD values reported in this study follow CLSI EP17-A2 verification criteria with ≥95% detection in 20 replicates, while regression-based estimates are lower but less reliable. It is therefore likely that the real analytical sensitivity of our assay may even be more superior, but we here report a more conservative value. While the ability to detect ultra-low viral loads could be advantageous in early infection or subclinical cases^31^, we acknowledge that its clinical significance should be interpreted with caution, particularly in distinguishing active infection from non-infectious shedding or residual DNA.

This study establishes the analytical performance of a direct real-time PCR assay, including in silico inclusivity and specificity analyses, in vitro determination of limit of detection, and verification of analytical sensitivity in spiked clinical matrix. It is not a diagnostic accuracy study conducted on naturally infected clinical specimens with an independent clinical reference method. Accordingly, prevalence-dependent performance metrics such as positive predictive value (PPV), negative predictive value (NPV), and agreement statistics such as kappa coefficient were not evaluated. Such indices require case-based sampling and comparison against a clinical reference standard, which falls beyond the analytical validation framework and objectives of the present study.

Nonetheless, we believe that the robust sensitivity and specificity of our direct, multiplex, clade-specific real-time PCR assay, that delivers results in under 1 hour, is a promising diagnostic tool for use in both surveillance and outbreak settings. Our methodological approach is comparable to many other studies^31,34–36^, which offers important insight and information as the first key step to advance diagnostics in an emerging disease. Further validation using naturally infected clinical specimens and real-world implementation studies will be essential to fully establish its clinical utility and operational feasibility.

## Competing interests statement

T.H.H. is a scientific advisor and H.C., B.L.H.K., and M.-S.T. are employees of ZYTCA Limited. Their conflicts of interest have been declared in the ICJME forms.

## Data availability statement

All sequences and data pertaining to this study can either be found in Supplementary Information or public databases. Accession numbers have been included in Supplementary Information.

## Ethics approval statement

This study was approved by the Institutional Review Board of Shanghai Children’s Medical Center (SCMCIRB-K2025018-1).

## Author contributions

H.C.: conceptualisation, investigation, validation, project administration, supervision, writing – review & editing;

T.H.H.: conceptualisation, formal analysis, investigation, methodology, validation, visualisation, writing – original draft;

B.L.: data curation, formal analysis, investigation, writing – review & editing;

B.L.H.K.: conceptualisation, data curation, formal analysis, investigation, methodology, writing – review & editing

Y.W.: data curation, investigation, writing – review & editing;

M.-S.T.: investigation, project administration, supervision, validation, writing – review & editing

D.D.: data curation, funding acquisition, investigation, resources, writing – review & editing

Z.C.: conceptualisation, project administration, supervision, writing – review & editing

## Acknowledgements

D.D. is supported by the National Natural Science Foundation of China (ref. 82173396).

## Notes

### Competing Interest Statement

T.H.H. is a scientific advisor and H.C., B.L.H.K., A.A.M., and M.-S.T. are employees of ZYTCA Limited.

### Author Declarations

The Institutional Review Board of Shanghai Children's Medical Center, School of Medicine, Shanghai Jao Tong University gave ethical approval for this work. Reference number is SCMCIRB-K2025018-1.

### Summary of Updates

A complementary probit curve analysis conducted; Revised Method, Results, and Discussion accordingly; Supplemental Figure 1 added.

