## Supplemental Information for "Evaluation and clinical validation of pan-specific and clade-specific diagnostic real-time PCR assays for monkeypox virus"

Hong Chang, PhD^1,2,^^, Tin Hang Hung, DPhil^3,^^
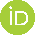
<https://orcid.org/0000-0001-9853-2053>, Binbin Li, MVM^1^, Bobby Lim Ho Kong, PhD^2^
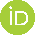
<https://orcid.org/0000-0001-9626-3128>, Ashwathi Asha Madhaven, PhD^2^, Yue Wang, BSc^1^, Ming-shan Tsai, DPhil^2^
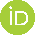
<https://orcid.org/0000-0001-6804-1317>, Dan Deng, MD, PhD^4^
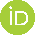
<https://orcid.org/0000-0001-7317-283X>, Zhanfeng Cui, PhD, DSc^1,5,*^

1. Oxford Suzhou Centre for Advanced Research (OSCAR), University of Oxford, Suzhou Industrial Park, Jiangsu, 215123, China
2. ZYTCA Limited, Centre for Innovation and Enterprise, Begbroke Science Park, Oxford, OX5 1PF, United Kingdom
3. Department of Biology, University of Oxford, OX1 3RB, United Kingdom
4. Department of Dermatology, Shanghai Children's Medical Center, School of Medicine, Shanghai Jiao Tong University, Shanghai 200127, China
5. Institute of Biomedical Engineering, Department of Engineering Science, University of Oxford, OX3 7DQ, United Kingdom

^^^H.C. and T.H.H. contributed equally as co-first authors.

Corresponding author:

*Z.C.:

**Supplementary Table 1.** List of 40 non-target high priority organisms from the WHO instructions and requirements for emergency use listing submission on in vitro diagnostic testing of MPXV nucleic acid (PQDx_457 version).

| **Family** | **Species** | **NCBI representative genome accession** | **NCBI Taxonomy ID** |
| --- | --- | --- | --- |
| Poxviridae | Buffalopox virus | OK422495 | 32605 |
| Poxviridae | Camelpox virus | NC_003391 | 28873 |
| Poxviridae | Cowpox virus | NC_003663 | 10243 |
| Poxviridae | Ectromelia (mousepox) virus | NC_004105 | 12643 |
| Poxviridae | Molluscum contagiosum virus | NC_001731 | 10279 |
| Poxviridae | Bovine papular stomatitis virus | NC_005337 | 129727 |
| Poxviridae | Orf virus | NC_005336 | 10258 |
| Poxviridae | Parapoxvirus | NC_025963 | 10257 |
| Poxviridae | Pseudocowpox virus | NC_013804 | 129726 |
| Poxviridae | Seal parapoxvirus | NC_035188 | 187984 |
| Poxviridae | Variola virus | LT706529 | 10255 |
| Poxviridae | Vaccinia virus | NC_006998 | 10245 |
| Non-Poxviridae | Herpes simplex virus 1 (HSV-1) | NC_001806 | 10298 |
| Non-Poxviridae | Herpes simplex virus 2 (HSV-2) | NC_001798 | 10310 |
| Non-Poxviridae | Varicella-zoster (chickenpox) virus | NC_001348 | 10335 |
| Non-Poxviridae | Streptococcus mitis | NZ_CP012646 | 28037 |
| Non-Poxviridae | Staphylococcus aureus | NC_007795 | 1280 |
| Non-Poxviridae | Staphylococcus epidermidis | NZ_CP035288 | 1282 |
| Non-Poxviridae | Streptococcus pyogenes | NZ_LS483338 | 1314 |
| Non-Poxviridae | Streptococcus agalactiae | NZ_CP012480 | 1311 |
| Non-Poxviridae | Pseudomonas aeruginosa | NC_002516 | 287 |
| Non-Poxviridae | Trichophyton rubrum | Multiple | 5551 |
| Non-Poxviridae | Corynebacterium jeikeium | NZ_CP063195 | 38289 |
| Non-Poxviridae | Candida albicans | Multiple | 5476 |
| Non-Poxviridae | Human Genomic DNA | Multiple | 9606 |
| Non-Poxviridae | Lactobacillus species (crispatus) | NZ_CP039266 | 47770 |
| Non-Poxviridae | Escherichia coli | NC_000913 | 562 |
| Non-Poxviridae | Acinetobacter calcoaceticus | NZ_CP020000 | 471 |
| Non-Poxviridae | Bacteroides fragilis | NC_003228 | 817 |
| Non-Poxviridae | Enterococcus faecalis | NZ_CP118962 | 1351 |
| Non-Poxviridae | Streptococcus Group C | AP012976 | 33972 |
| Non-Poxviridae | Streptococcus Group G | Multiple | 1320 |
| Non-Poxviridae | Corynebacterium diphtheriae | NZ_LN831026 | 1717 |
| Non-Poxviridae | Neisseria gonorrhoeae | NZ_AP023069 | 485 |
| Non-Poxviridae | Chlamydia trachomatis | NC_000117 | 813 |
| Non-Poxviridae | Mycoplasmoides pneumoniae | NZ_LR214945 | 2104 |
| Non-Poxviridae | Mycoplasmoides genitalium | NC_000908 | 2097 |
| Non-Poxviridae | Human papilloma virus (HPV) | NC_027779 | 10566 |
| Non-Poxviridae | Trichomonas vaginalis | Multiple | 5722 |
| Non-Poxviridae | Treponema pallidum | NC_021490 | 160 |

**Supplementary Table 2.** Sequences and statistics for all primers and probes listed in this study.

|  | **Sequence** | **Amplicon** | **Length** |
| --- | --- | --- | --- |
| Mpox_F3L_FP | CATCTATTATAGCATCAGCATCAGA | catctattatagcatcagcatcagaatctgtaggccgtgtatcagcatccattgtcgtagaccaacgaggaggagtatc | 79 |
| Mpox_F3L_RP | GATACTCCTCCTCGTTGGTCTAC |  |  |
| Mpox_F3L_Probe | TGTAGGCCGTGTATCAGCATCCATT |  |  |
| Mpox_B15L_FP | ACTAACTGGTCAGCAGGCCATT | actaactggtcagcaggccattagccaacagactagatcaactacgttgagtagaaaagatcagatgagcaaggaagaaaagatattcgaagcagttacaatgagtctatcaactataggttcaacgttgacgtctgcaggtatgacgggtggtccaaaactaatgattgcaggaatggctataacggct | 190 |
| Mpox_B15L_RP | AGCCGTTATAGCCATTCCTGCA |  |  |
| Mpox_B15L_Probe | TGGACCACCCGTCATRCCTGCAGAC |  |  |
| Mpox_B1R_FP | CATGGATTTTTGATGGTGGTTTAAG | catggatttttgatggtggtttaagtttaaaaaagattttgttattgtagtatgataatatcaaaaagatggatataaagaattggtcagtgtataataaattatatgtaggaggaggaatatctgatgatgttc | 135 |
| Mpox_B1R_RPv1 | GAACATCATCAGATATTCCTCCTCC |  |  |
| Mpox_B1R_Probe | AAGATGGATATAAAGAATTGGTCAGTGT |  |  |

**Supplementary Table 3.** List of 56 MPXV genomes used for multiple genome alignment and clade-specific primer design.

| **Accession** | **Country*** | **Year** | **Clade** |
| --- | --- | --- | --- |
| NC_003310 | DRC | 1996 | I |
| NC_063383 | Nigeria | 2020 | II |
| KJ642618 | Cameroon | 1989 | I |
| KJ642619 | Gabon | 1987 | I |
| OP498046 | Gabon | 1988 | I |
| KJ642613 | DRC | 1970 | I |
| DQ011154 | DRC | 2003 | I |
| MN702448 | CAR | 2018 | I |
| MN702453 | CAR | 2001 | I |
| MN702451 | CAR | 2017 | I |
| MN702444 | CAR | 2018 | I |
| KP849471 | DRC | 1985 | I |
| JX878411 | DRC | 2006 | I |
| JX878422 | DRC | 2007 | I |
| JX878420 | DRC | 2007 | I |
| KJ642612 | DRC | 1986 | I |
| HM172544 | DRC | 1979 | I |
| U84504 | Gabon | 1998 | I |
| JX878417 | DRC | 2007 | I |
| OP580587 | USA | 2022 | II |
| KJ642617 | USA | 1971 | II |
| MG693724 | Nigeria | 2017 | II |
| OP587264 | USA | 2022 | II |
| DQ011156 | Liberia | 1970 | II |
| KP849470 | Cote d'Ivoire | 1970 | II |
| AY741551 | Sierra Leone | 1970 | II |
| AY603973 | USA | 1961 | II |
| DQ011157 | USA | 2003 | II |
| KJ642616 | France | 1968 | II |
| KJ642614 | Netherlands | 1965 | II |
| AY753185 | Denmark | 1958 | II |
| KJ642615 | Nigeria | 1978 | II |
| KJ136820 | Cote d'Ivoire | 2012 | II |
| OP764629 | Germany | 2022 | II |
| OP764614 | Germany | 2022 | II |
| OP752116 | USA | 2022 | II |
| MT903338 | Nigeria | 2017 | II |
| MT903339 | Nigeria | 2018 | II |
| MT903342 | Singapore | 2019 | II |
| MT903344 | UK | 2018 | II |
| ON595760 | Switzerland | 2022 | II |
| ON614676 | Italy | 2022 | II |
| ON615424 | Netherlands | 2022 | II |
| ON619837 | UK | 2022 | II |
| ON622712 | Belgium | 2022 | II |
| ON622722 | France | 2022 | II |
| ON631963 | Australia | 2022 | II |
| ON649710 | Portugal | 2022 | II |
| ON674051 | USA | 2022 | II |
| ON675438 | USA | 2022 | II |
| ON676706 | USA | 2022 | II |
| ON676707 | USA | 2021 | II |
| ON676708 | USA | 2021 | II |
| ON682264 | Germany | 2022 | II |
| ON736420 | Canada | 2022 | II |
| MN648051 | Israel | 2018 | II |

**Supplementary Table 4.** List of randomly selected representative genomes for each organism listed in Supplementary Table 1 for cross-reactivity analysis.

| **Accession** | **Species** | **NCBI Taxonomy ID** |
| --- | --- | --- |
| GCA_006451575.1 | Buffalopox virus | 32605 |
| GCA_013387605.1 | Buffalopox virus | 32605 |
| GCA_013387595.1 | Buffalopox virus | 32605 |
| GCA_021870145.1 | Buffalopox virus | 32605 |
| GCA_013387585.1 | Buffalopox virus | 32605 |
| GCA_020809605.1 | Camelpox virus | 28873 |
| GCA_006458065.1 | Camelpox virus | 28873 |
| GCA_020809645.1 | Camelpox virus | 28873 |
| GCA_030247805.1 | Camelpox virus | 28873 |
| GCA_020809635.1 | Camelpox virus | 28873 |
| GCA_023530455.1 | Cowpox virus | 10243 |
| GCA_023533615.1 | Cowpox virus | 10243 |
| GCA_023535875.1 | Cowpox virus | 10243 |
| GCA_900187825.1 | Cowpox virus | 10243 |
| GCA_023535735.1 | Cowpox virus | 10243 |
| GCA_023531875.1 | Ectromelia virus | 12643 |
| GCA_031198905.1 | Ectromelia virus WH | 12643 |
| GCA_006458505.1 | Ectromelia virus ERPV | 12643 |
| GCA_006465865.1 | Ectromelia virus Naval | 12643 |
| GCF_000841905.1 | Ectromelia virus | 12643 |
| GCA_016865705.1 | Molluscum contagiosum virus | 10279 |
| GCA_035088935.1 | Molluscum contagiosum virus | 10279 |
| GCA_016865665.1 | Molluscum contagiosum virus | 10279 |
| GCA_035089025.1 | Molluscum contagiosum virus | 10279 |
| GCA_035089015.1 | Molluscum contagiosum virus | 10279 |
| GCA_023536365.1 | Bovine papular stomatitis virus | 129727 |
| GCA_038370105.1 | Bovine papular stomatitis virus | 129727 |
| GCA_023536345.1 | Bovine papular stomatitis virus | 129727 |
| GCF_000844045.1 | Bovine papular stomatitis virus | 129727 |
| GCA_023536355.1 | Bovine papular stomatitis virus | 129727 |
| GCA_023531965.1 | Orf virus | 10258 |
| GCA_023536275.1 | Orf virus | 10258 |
| GCA_023533605.1 | Orf virus | 10258 |
| GCA_024426035.1 | Orf virus | 10258 |
| GCA_023536405.1 | Orf virus | 10258 |
| GCA_023536355.1 | Bovine papular stomatitis virus | 10257 |
| GCA_023536275.1 | Orf virus | 10257 |
| GCA_023536345.1 | Bovine papular stomatitis virus | 10257 |
| GCA_032918225.1 | Equine parapoxvirus | 10257 |
| GCA_023531815.1 | Orf virus | 10257 |
| GCA_038370045.1 | Pseudocowpox virus | 129726 |
| GCA_023533575.1 | Pseudocowpox virus | 129726 |
| GCF_000886295.1 | Pseudocowpox virus | 129726 |
| GCA_000886295.1 | Pseudocowpox virus | 129726 |
| GCA_038370085.1 | Pseudocowpox virus | 129726 |
| GCA_002219465.1 | Seal parapoxvirus | 187984 |
| GCF_002219465.1 | Seal parapoxvirus | 187984 |
| GCA_037113675.1 | Variola virus | 10255 |
| GCA_037114905.1 | Variola virus | 10255 |
| GCA_023530375.1 | Variola virus | 10255 |
| GCA_025630175.1 | Variola virus | 10255 |
| GCA_006465665.1 | Variola major virus | 10255 |
| GCA_023535885.1 | Vaccinia virus | 10245 |
| GCA_023533815.1 | Vaccinia virus | 10245 |
| GCA_038422455.1 | Modified Vaccinia Ankara virus | 10245 |
| GCA_023532155.1 | Vaccinia virus | 10245 |
| GCA_023532135.1 | Vaccinia virus | 10245 |
| GCA_027937575.1 | Human alphaherpesvirus 1 | 10298 |
| GCA_027937905.1 | Human alphaherpesvirus 1 | 10298 |
| GCA_027937355.1 | Human alphaherpesvirus 1 | 10298 |
| GCA_027937955.1 | Human alphaherpesvirus 1 | 10298 |
| GCA_027939335.1 | Human alphaherpesvirus 1 | 10298 |
| GCA_027936455.1 | Human alphaherpesvirus 2 | 10310 |
| GCA_003052145.1 | Human alphaherpesvirus 2 | 10310 |
| GCA_027937075.1 | Human alphaherpesvirus 2 | 10310 |
| GCA_027936265.1 | Human alphaherpesvirus 2 | 10310 |
| GCA_027937255.1 | Human alphaherpesvirus 2 | 10310 |
| GCA_027941095.1 | Human alphaherpesvirus 3 | 10335 |
| GCA_027941135.1 | Human alphaherpesvirus 3 | 10335 |
| GCA_027941165.1 | Human alphaherpesvirus 3 | 10335 |
| GCF_000858285.1 | Human alphaherpesvirus 3 | 10335 |
| GCA_027941045.1 | Human alphaherpesvirus 3 | 10335 |
| GCF_000722765.2 | Streptococcus mitis | 28037 |
| GCF_016658865.1 | Streptococcus mitis | 28037 |
| GCF_000027165.1 | Streptococcus mitis B6 | 28037 |
| GCA_000027165.1 | Streptococcus mitis B6 | 28037 |
| GCA_019047825.1 | Streptococcus mitis | 28037 |
| GCF_029094305.1 | Staphylococcus aureus | 1280 |
| GCA_009913215.1 | Staphylococcus aureus | 1280 |
| GCA_014725655.1 | Staphylococcus aureus | 1280 |
| GCF_900635315.1 | Staphylococcus aureus | 1280 |
| GCF_045346715.1 | Staphylococcus aureus | 1280 |
| GCA_003856455.1 | Staphylococcus epidermidis | 1282 |
| GCA_022832935.1 | Staphylococcus epidermidis | 1282 |
| GCA_042464785.1 | Staphylococcus epidermidis | 1282 |
| GCA_045935655.1 | Staphylococcus epidermidis | 1282 |
| GCF_029457575.1 | Staphylococcus epidermidis | 1282 |
| GCA_045290705.1 | Streptococcus pyogenes | 1314 |
| GCA_002557755.1 | Streptococcus pyogenes | 1314 |
| GCF_000230295.1 | Streptococcus pyogenes Alab49 | 1314 |
| GCF_000993765.1 | Streptococcus pyogenes | 1314 |
| GCF_034434355.2 | Streptococcus pyogenes | 1314 |
| GCA_949788945.1 | Streptococcus agalactiae | 1311 |
| GCA_030908345.1 | Streptococcus agalactiae | 1311 |
| GCA_034329605.1 | Streptococcus agalactiae | 1311 |
| GCF_001592425.1 | Streptococcus agalactiae | 1311 |
| GCA_002289205.1 | Streptococcus agalactiae | 1311 |
| GCF_044757545.1 | Pseudomonas aeruginosa | 287 |
| GCA_027570775.1 | Pseudomonas aeruginosa | 287 |
| GCA_033971305.1 | Pseudomonas aeruginosa | 287 |
| GCF_023101265.1 | Pseudomonas aeruginosa | 287 |
| GCF_024734815.1 | Pseudomonas aeruginosa | 287 |
| GCA_002209185.2 | Corynebacterium jeikeium | 38289 |
| GCF_028609885.1 | Corynebacterium jeikeium | 38289 |
| GCF_000006605.1 | Corynebacterium jeikeium K411 | 38289 |
| GCA_028609885.1 | Corynebacterium jeikeium | 38289 |
| GCA_003955985.1 | Corynebacterium jeikeium | 38289 |
| GCA_032688725.1 | Candida albicans | 5476 |
| GCA_041438225.1 | Candida albicans | 5476 |
| GCA_964212385.1 | Homo sapiens | 9606 |
| GCA_964212445.1 | Homo sapiens | 9606 |
| GCA_964212355.1 | Homo sapiens | 9606 |
| GCA_964212285.1 | Homo sapiens | 9606 |
| GCA_964212455.1 | Homo sapiens | 9606 |
| GCF_029011475.1 | Lactobacillus crispatus | 47770 |
| GCA_041344855.1 | Lactobacillus crispatus | 47770 |
| GCA_009933525.1 | Lactobacillus crispatus | 47770 |
| GCF_013456995.1 | Lactobacillus crispatus | 47770 |
| GCF_000466885.3 | Lactobacillus crispatus 2029 | 47770 |
| GCA_043987575.1 | Escherichia coli | 562 |
| GCA_043944135.1 | Escherichia coli | 562 |
| GCA_030389595.1 | Escherichia coli | 562 |
| GCF_037553545.1 | Escherichia coli | 562 |
| GCA_002156825.1 | Escherichia coli | 562 |
| GCF_016939475.1 | Acinetobacter calcoaceticus | 471 |
| GCF_900520355.1 | Acinetobacter calcoaceticus | 471 |
| GCA_021165835.1 | Acinetobacter calcoaceticus | 471 |
| GCA_039725545.1 | Acinetobacter calcoaceticus | 471 |
| GCA_032594495.1 | Acinetobacter calcoaceticus | 471 |
| GCF_021359565.1 | Bacteroides fragilis | 817 |
| GCF_024759945.1 | Bacteroides fragilis | 817 |
| GCA_024759945.1 | Bacteroides fragilis | 817 |
| GCF_000710375.2 | Bacteroides fragilis | 817 |
| GCA_024758125.1 | Bacteroides fragilis | 817 |
| GCA_905123065.1 | Enterococcus faecalis | 1351 |
| GCF_036277495.1 | Enterococcus faecalis | 1351 |
| GCA_000172575.2 | Enterococcus faecalis OG1RF | 1351 |
| GCF_905123245.1 | Enterococcus faecalis | 1351 |
| GCF_028335505.1 | Enterococcus faecalis | 1351 |
| GCA_004758745.1 | Corynebacterium diphtheriae | 1717 |
| GCF_900638705.1 | Corynebacterium diphtheriae | 1717 |
| GCF_002843135.1 | Corynebacterium diphtheriae bv. mitis str. ISS 3319 | 1717 |
| GCA_021560315.1 | Corynebacterium diphtheriae | 1717 |
| GCF_963556575.1 | Corynebacterium diphtheriae | 1717 |
| GCA_025790185.1 | Neisseria gonorrhoeae | 485 |
| GCF_900087815.2 | Neisseria gonorrhoeae | 485 |
| GCA_023611785.1 | Neisseria gonorrhoeae | 485 |
| GCA_025642335.1 | Neisseria gonorrhoeae | 485 |
| GCA_025170665.2 | Neisseria gonorrhoeae | 485 |
| GCA_021049405.1 | Chlamydia trachomatis | 813 |
| GCF_000319105.1 | Chlamydia trachomatis L3/404/LN | 813 |
| GCF_021047725.1 | Chlamydia trachomatis | 813 |
| GCF_000507225.1 | Chlamydia trachomatis C/TW-3 | 813 |
| GCF_021050805.1 | Chlamydia trachomatis | 813 |
| GCF_001509195.1 | Mycoplasmoides pneumoniae | 2104 |
| GCA_000319655.2 | Mycoplasmoides pneumoniae PO1 | 2104 |
| GCA_009940325.1 | Mycoplasmoides pneumoniae | 2104 |
| GCA_002128065.1 | Mycoplasmoides pneumoniae | 2104 |
| GCF_001272835.1 | Mycoplasmoides pneumoniae FH | 2104 |
| GCF_036784995.1 | Mycoplasmoides genitalium | 2097 |
| GCA_000292445.1 | Mycoplasmoides genitalium M6282 | 2097 |
| GCA_000292405.1 | Mycoplasmoides genitalium M2321 | 2097 |
| GCA_036784995.1 | Mycoplasmoides genitalium | 2097 |
| GCF_000027325.1 | Mycoplasmoides genitalium G37 | 2097 |
| GCA_001274345.1 | Human papillomavirus | 10566 |
| GCF_001274345.1 | Human papillomavirus | 10566 |
| GCF_023016365.1 | Treponema pallidum subsp. pallidum | 160 |
| GCA_029866705.1 | Treponema pallidum subsp. pertenue | 160 |
| GCF_023016385.1 | Treponema pallidum subsp. pallidum | 160 |
| GCF_023016445.1 | Treponema pallidum subsp. pallidum | 160 |
| GCF_023016425.1 | Treponema pallidum subsp. pallidum | 160 |

**Supplementary Table 5.** Sequences of **(a)** Clade I and **(b)** Clade II MPXV synthetic standards. Red, blue, and green sequences denote the F3L, B15L, and B1R amplicons.

**(a)**

GGCTGTTGTCCTGCATCGGAACCTCACTCATGCATCTCATGCCTCCTATCACGCTCTCGAGTGACTCATCTATTATAGCATCAGCATCAGAATCTGTAGGCCGTGTATCAGCATCCATTGTCGTAGACCAACGAGGAGGAGTATCCTGCCGAGAGCCGGCCATTGTCATTGGGGCCTCACACTAACTGGTCAGCAGGCCATTAGCCAACAGACTAGATCAACTACGTTGAGTAGAAAAGATCAGATGAGCAAGGAAGAAAAGATATTCGAAGCAGTTACAATGAGTCTATCAACTATAGGTTCAACGTTGACGTCTGCAGGTATGACGGGTGGTCCAAAACTAATGATTGCAGGAATGGCTATAACGGCTCCGGCAAGCAAGCATGAACAACGCAAGGATCGGCGAGTTGTGGATGTTCGGGTTCGACATCCACCGATGGTGTCACGCCACTAATCGGTTCGGTAACGTCTGTGGATGGAGGTGCTACTTAGGTCGGGCAGGGAATTAAGGGGAACGCATGCTCGGCGCAATGAAGCTGGGGGACCGGCGTGGGGACCGCGCCCCTGCCTAAGCGAGAATCATGGATTTTTGATGGTGGTTTAAGTTTAAAAAAGATTTTGTTATTGTAGTATGATAATATCAAAAAGATGGATATAAAGAATTGGTCAGTGTATAATAAATTATATGTAGGAGGAGGAATATCTGATGATGTTCAAACTAATACATCTGAAACATACGATAAAGAAAAAGATTGTTGGACATTGGATAATGGTCACTTGGTACCACGGGCACGGGGTGGCAACCGCCCCCCTGGCGTGCGGGGCGCACTCCGTAACGATTTGGGGGTCCGGAGACTCGCCGCTTCCGGGATTCCCC

**(b)**

TCATTATTCCTATCACGCTTTCGAGTGACTCATCTATTATAGCATCAGCATCAGAATCTGTAGGCCGTGTATCAGCATCCATTGTCGTAGACCAACGAGGAGGAGTATCCTGCCGAGAGCCGGCCATTGTCATTGGGGCCTCACACTAACTGGTCAGCAGGCCATTAGCCAACAGACTAGATCAACTACGTTGAGTAGAAAAGATCAGATGAGCAAGGAAGAAAAGATATTCGAAGCAGTTACAATGAGTCTATCAACTATAGGTTCAACGTTGACGTCTGCAGGTATGACGGGTGGTCCAAAACTAATGATTGCAGGAATGGCTATAACGGCTCCGGCAAGCAATTATGAACAACGCAAGGATCGGCGAGTTGTGGATGTTCGGGTTCGACATCCACCGATGGTGTCACGCCACTAATCGGTTCGGTAACGTCTGTGGATGGAGGTGCTACTTCAGATGCTGGCCACGAGCTAAATTATATCACTACGGACATAAACCATTGTATAATTTTTATGTTTATTAGTGTACACATTTTGGAAGTAAGTTCCTGGATCGGATGTCACCGCAGTAATATTGTTGATTCCCTATTGAGGCATTGACTGATGCGGGAAGAGATC

**Supplementary Table 6**. List of human skin lesion samples used for the MPXV clinical negative sample matrix.

| **Sample** | **Symptoms** | Gender |
| --- | --- | --- |
| 1 | Perianal abscess pus | Male |
| 2 | Forehead acne | Male |
| 3 | Forehead comedones | Female |
| 4 | Forehead comedones | Male |
| 5 | Low temperature burn blister fluid on the left lower limb | Female |
| 6 | Leg scabies pus | Male |
| 7 | Forehead comedones | Female |
| 8 | Femoral tinea | Male |
| 9 | Palm desquamation | Male |
| 10 | Left ankle eczema scales | Male |
| 11 | Nasal pus | Male |
| 12 | Acne | Female |
| 13 | Temporal abscess pus | Male |
| 14 | Abscess scabies | Male |
| 15 | Dermatitis | Female |
| 16 | Skin infection | Male |
| 17 | Burn | Male |
| 18 | Diaper rash | Male |
| 19 | Acne | Male |
| 20 | Right thigh dermatitis | Female |
| 21 | Eczema | Male |
| 22 | Dermatitis | Female |
| 23 | Tinea pedis | Female |
| 24 | Acne | Male |
| 25 | Acne | Male |
| 26 | Pompholyx | Female |
| 27 | Calf eczema | Male |
| 28 | Right breast skin scabies | Female |
| 29 | Paronychial abscess | Female |
| 30 | Diaper rash | Male |
| 31 | Facial tinea | Female |
| 32 | Acne | Female |
| 33 | Keratolysis exfoliativa | Male |
| 34 | Dandruff | Male |
| 35 | Diaper rash | Male |
| 36 | Seborrheic dermatitis of the scalp | Female |
| 37 | Acne | Male |
| 38 | Acne | Male |
| 39 | Tinea pedis | Female |
| 40 | Seborrheic dermatitis of the scalp | Male |
| 41 | Eczema | Female |
| 42 | Verruca vulgaris | Male |
| 43 | Diaper rash | Male |
| 44 | Hand tinea | Male |
| 45 | Back eczema | Male |
| 46 | Palm keratolysis exfoliativa | Male |
| 47 | Tinea pedis | Female |
| 48 | Eczema with infection | Male |
| 49 | Athlete's foot | Female |
| 50 | Tinea pedis with infection | Male |

**Supplementary Table 7**. Statistics of potential off-target amplification from two cowpox virus genomes.

| **Potential off-target** | **Match** | **Identity (%)** | **Length** | **Mismatch** | **Start of sequence** | **End of sequence** | **Start of match** | **End of match** | **E-value** | **Bitscore** |
| --- | --- | --- | --- | --- | --- | --- | --- | --- | --- | --- |
| GCA_023530455.1 | B15L_FP | 100 | 22 | 0 | 207559 | 207580 | 1 | 22 | 2.06E-06 | 44.1 |
|  | B15L_Probe | 92 | 25 | 2 | 207691 | 207715 | 25 | 1 | 0.000127 | 38.2 |
|  | B15L_RP | 95.455 | 22 | 1 | 207727 | 207748 | 22 | 1 | 0.000501 | 36.2 |
| GCA_023533615.1 | B15L_FP | 100 | 22 | 0 | 205942 | 205963 | 1 | 22 | 2.04E-06 | 44.1 |
|  | B15L_Probe | 88 | 25 | 3 | 206074 | 206098 | 25 | 1 | 0.031 | 30.2 |
|  | B15L_RP | 100 | 22 | 0 | 206110 | 206131 | 22 | 1 | 2.04E-06 | 44.1 |

**Supplementary Table 8**. Ct values from real-time PCR for determination of standard curve for MPXV synthetic standards.

| **Copy number** | **Ct value** | |
| --- | --- | --- |
|  | **Pan-specific (B15L+F3L)** | **Clade I-specific (B1R)** |
| 2.00E+06 | 16.547 | 17.438 |
|  | 16.534 | 17.442 |
|  | 16.537 | 17.414 |
| 2.00E+05 | 19.906 | 20.762 |
|  | 19.967 | 20.762 |
|  | 19.839 | 20.616 |
| 2.00E+04 | 23.235 | 23.956 |
|  | 23.165 | 23.908 |
|  | 23.295 | 24.002 |
| 2.00E+03 | 26.758 | 27.408 |
|  | 26.667 | 27.304 |
|  | 26.86 | 27.461 |
| 2.00E+02 | 30.264 | 30.831 |
|  | 30.366 | 30.934 |
|  | 30.186 | 30.793 |
| 2.00E+01 | 33.634 | 34.058 |
|  | 33.855 | 34.164 |
|  | 33.688 | 34.253 |
| 2.00E+00 | 34.323 | 34.508 |
|  | 34.482 | 34.602 |
|  | 34.740 | 35.118 |
| NC | - | - |
|  | - | - |
|  | - | - |

**Supplementary Table 9**. Ct values from real-time PCR for tentative limit of detection of MPXV synthetic standard. NC denotes negative control.

| **Copy number** | **Ct value** | | |
| --- | --- | --- | --- |
|  | **Pan-specific** | | **Clade I-specific** |
|  | **B15L** | **F3L** | **B1R** |
| 20,000 | 24.205 | 24.118 | 24.250 |
|  | 24.108 | 23.954 | 24.259 |
|  | 24.209 | 24.209 | 24.270 |
| 2,000 | 27.641 | 27.446 | 27.574 |
|  | 27.703 | 27.648 | 27.688 |
|  | 27.463 | 27.576 | 27.568 |
| 200 | 31.095 | 30.739 | 30.931 |
|  | 30.863 | 30.925 | 30.947 |
|  | 30.779 | 30.674 | 31.166 |
| 20 | 34.088 | 34.534 | 34.145 |
|  | 34.022 | 34.323 | 33.989 |
|  | 33.745 | 34.266 | 34.390 |
| 2 | 36.956 | 36.292 | 37.950 |
|  | 38.041 | 38.345 | 39.160 |
|  | 37.109 | 37.527 | 37.624 |
| 0.2 | 37.151 | - | - |
|  | 37.143 | - | - |
|  | - | 39.114 | - |
| NC | - | - | - |
|  | - | - | - |
|  | - | - | - |

**Supplementary Table 10.** Ct values from real-time PCR for analytical limit of detection using 2 copies of MPXV synthetic standard per reaction. NC denotes negative control.

| **Replicate** | **Ct value** | | |
| --- | --- | --- | --- |
|  | **Pan-specfic** | | **Clade I-specific** |
|  | **B15L** | **F3L** | **B1R** |
| 1 | 36.647 | 35.952 | 37.577 |
| 2 | 35.93 | 36.784 | 37.328 |
| 3 | 35.949 | 35.662 | 36.450 |
| 4 | 35.595 | 36.675 | 36.099 |
| 5 | 34.753 | 35.875 | 37.308 |
| 6 | 35.994 | 35.337 | 38.107 |
| 7 | 36.704 | 36.076 | 36.533 |
| 8 | 36.607 | 36.471 | 36.963 |
| 9 | 35.763 | 35.593 | 36.305 |
| 10 | 36.097 | 37.853 | 37.573 |
| 11 | 35.636 | 35.953 | 36.400 |
| 12 | 35.546 | 35.447 | 37.455 |
| 13 | 35.961 | 35.173 | 36.934 |
| 14 | 37.253 | 35.892 | 37.519 |
| 15 | 36.122 | 35.117 | 36.869 |
| 16 | 36.926 | 35.248 | 36.999 |
| 17 | 36.907 | 35.159 | 36.364 |
| 18 | 35.656 | 35.666 | 36.413 |
| 19 | 37.515 | 36.695 | 36.521 |
| 20 | 36.059 | 35.237 | 37.675 |
| NC1 | - | - | - |
| NC2 | - | - | - |
| NC3 | - | - | - |

**Supplementary Table 11.** Ct values from real-time PCR for tentative limit of detection of MPXV Clade I synthetic standard spiked in clinical negative matrix. NC denotes negative control.

| **Copy number**  **/reaction** | **Batch 1** | | | | **Batch 2** | | | |
| --- | --- | --- | --- | --- | --- | --- | --- | --- |
|  | **Tube A** | | **Tube B** | | **Tube A** | | **Tube B** | |
|  | **FAM**  **B15L+F3L** | **HEX**  **B1R** | **FAM**  **SPC** | **HEX**  **ACTB** | **FAM**  **B15L+F3L** | **HEX**  **B1R** | **FAM**  **SPC** | **HEX**  **ACTB** |
| 3.0 | 33.81 | 37.18 | 31.98 | 29.86 | 33.53 | 36.66 | 32.16 | 29.46 |
|  | 33.40 | 36.32 | 32.06 | 29.88 | 33.47 | 36.68 | 32.04 | 29.58 |
|  | 33.42 | 36.69 | 32.13 | 29.79 | 33.96 | 38.29 | 32.22 | 29.75 |
| 2.5 | 33.87 | 37.60 | 31.86 | 29.87 | 33.16 | 36.81 | 31.99 | 29.71 |
|  | 33.56 | 37.63 | 31.98 | 29.33 | 33.74 | 37.65 | 31.90 | 29.15 |
|  | 33.85 | 38.13 | 31.98 | 29.62 | 33.28 | 36.10 | 31.98 | 29.41 |
| 2.0 | 33.96 | 37.54 | 31.75 | 30.00 | 34.19 | 37.93 | 32.03 | 28.5 |
|  | 34.04 | 38.10 | 32.10 | 29.94 | 33.96 | 37.58 | 32.23 | 29.53 |
|  | 34.00 | 38.14 | 31.88 | 29.82 | 34.12 | 38.49 | 32.38 | 29.58 |
| 1.5 | 34.40 | - | 31.97 | 29.97 | 34.44 | 39.07 | 32.73 | 30.06 |
|  | 33.62 | 37.50 | 32.07 | 29.81 | 34.38 | - | 32.03 | 29.88 |
|  | 34.07 | 38.67 | 32.12 | 29.84 | 33.95 | 37.70 | 32.09 | 29.69 |
| 1.0 | 34.95 | 39.69 | 31.82 | 29.57 | 34.81 | - | 31.94 | 29.51 |
|  | 35.28 | 39.83 | 32.05 | 29.83 | 35.36 | - | 31.72 | 29.73 |
|  | 34.95 | - | 31.85 | 29.16 | 35.26 | - | 32.12 | 29.90 |
| 0 | - | - | 31.89 | 28.88 | - | - | 31.96 | 29.96 |
|  | - | - | 31.86 | 29.41 | - | - | 31.72 | 29.71 |
|  | - | - | 32.05 | 30.00 | - | - | 31.96 | 29.57 |
| NC | - | - | - | - | - | - | - | - |
|  | - | - | - | - | - | - | - | - |
|  | - | - | - | - | - | - | - | - |

**Supplementary Table 12.** Ct values from real-time PCR for analytical limit of detection of MPXV Clade I synthetic standard spiked in clinical negative matrix. NC denotes negative control.

| **Replicate** | **Batch 1** | | | | **Batch 2** | | | |
| --- | --- | --- | --- | --- | --- | --- | --- | --- |
|  | **Tube A** | | **Tube B** | | **Tube A** | | **Tube B** | |
|  | **FAM**  **B15L+F3L** | **HEX**  **B1R** | **FAM**  **SPC** | **HEX**  **ACTB** | **FAM**  **B15L+F3L** | **HEX**  **B1R** | **FAM**  **SPC** | **HEX**  **ACTB** |
| 1 | 34.25 | 36.37 | 32.91 | 30.16 | 34.53 | 37.15 | 33.37 | 29.86 |
| 2 | 34.85 | 36.85 | 33.08 | 30.31 | 34.01 | 35.78 | 32.65 | 29.80 |
| 3 | 34.66 | 36.58 | 32.91 | 30.12 | 35.21 | 38.23 | 33.02 | 29.99 |
| 4 | 35.08 | 37.03 | 33.02 | 30.17 | 34.74 | 37.43 | 32.47 | 29.66 |
| 5 | 34.56 | 36.67 | 32.68 | 30.22 | 34.32 | 36.51 | 32.92 | 30.24 |
| 6 | 35.01 | 37.21 | 33.21 | 30.12 | 35.27 | 37.89 | 33.06 | 29.83 |
| 7 | 33.14 | 34.74 | 33.23 | 30.41 | 34.3 | 36.33 | 32.95 | 29.58 |
| 8 | 34.84 | 36.76 | 32.85 | 30.14 | 34.96 | 37.59 | 32.6 | 29.66 |
| 9 | 34.78 | 36.86 | 32.66 | 30.21 | 34.5 | 36.92 | 32.48 | 29.61 |
| 10 | 34.72 | 36.79 | 32.54 | 30.35 | 34.49 | 36.85 | 32.08 | 30.06 |
| 11 | 34.88 | 36.63 | 32.92 | 29.95 | 34.26 | 36.68 | 32.85 | 29.88 |
| 12 | 34.21 | 36.26 | 32.74 | 30.10 | 34.75 | 37.26 | 32.69 | 29.59 |
| 13 | 35.39 | 37.73 | 32.95 | 30.12 | 34.41 | 36.47 | 32.87 | 29.79 |
| 14 | 35.44 | 39.16 | 32.47 | 30.22 | 34.02 | 36.27 | 32.68 | 29.69 |
| 15 | 34.96 | 37.16 | 32.83 | 30.44 | 34.54 | 36.97 | 32.78 | 29.88 |
| 16 | 34.79 | 37.09 | 32.66 | 30.07 | 34.81 | 37.38 | 33.37 | 29.89 |
| 17 | 34.60 | 36.59 | 33.06 | 30.11 | 34.09 | 36.08 | 32.97 | 30.06 |
| 18 | 35.28 | 37.58 | 32.47 | 30.16 | 34.44 | 36.73 | 32.53 | 30.00 |
| 19 | 35.11 | 37.28 | 32.47 | 29.84 | 34.87 | - | 32.88 | 30.1 |
| 20 | 34.66 | 36.92 | 32.91 | 30.48 | 34.59 | 36.72 | 32.91 | 29.5 |

**Supplementary Table 13.** Ct values from real-time PCR for tentative limit of detection of MPXV Clade II synthetic standard spiked in clinical negative matrix. NC denotes negative control.

| **Copy number**  **/reaction** | **Batch 1** | | | | **Batch 2** | | | |
| --- | --- | --- | --- | --- | --- | --- | --- | --- |
|  | **Tube A** | | **Tube B** | | **Tube A** | | **Tube B** | |
|  | **FAM**  **B15L+F3L** | **HEX**  **B1R** | **FAM**  **SPC** | **HEX**  **ACTB** | **FAM**  **B15L+F3L** | **HEX**  **B1R** | **FAM**  **SPC** | **HEX**  **ACTB** |
| 3.0 | 34.42 | - | 31.84 | 28.94 | 34.46 | - | 31.76 | 28.34 |
|  | 33.47 | - | 31.58 | 27.98 | 32.93 | - | 31.70 | 28.22 |
|  | 33.61 | - | 31.76 | 27.76 | 33.28 | - | 31.64 | 28.25 |
| 2.5 | 34.41 | - | 31.53 | 27.94 | 33.78 | - | 31.45 | 28.18 |
|  | 33.76 | - | 31.53 | 26.97 | 35.03 | - | 31.84 | 28.24 |
|  | 34.35 | - | 31.58 | 27.46 | 33.88 | - | 31.28 | 28.34 |
| 2.0 | 32.70 | - | 31.84 | 28.14 | 33.70 | - | 32.06 | 28.40 |
|  | 34.01 | - | 31.70 | 27.80 | 34.49 | - | 31.88 | 27.99 |
|  | 33.95 | - | 31.85 | 28.21 | 34.85 | - | 31.98 | 28.87 |
| 1.5 | 34.80 | - | 32.06 | 27.59 | 34.69 | - | 31.85 | 28.19 |
|  | 34.53 | - | 31.64 | 27.74 | 35.38 | - | 32.10 | 28.50 |
|  | 34.44 | - | 32.16 | 28.20 | 34.99 | - | 31.92 | 28.09 |
| 1.0 | 34.62 | - | 32.01 | 27.93 | 36.11 | - | 31.71 | 28.27 |
|  | 34.91 | - | 31.99 | 28.18 | 36.44 | - | 32.19 | 28.14 |
|  | 35.14 | - | 32.06 | 27.83 | 36.16 | - | 32.12 | 27.78 |
| 0 | - | - | 32.39 | 30.45 | - | - | 30.38 | 28.05 |
|  | - | - | 31.69 | 29.62 | - | - | 30.30 | 28.01 |
|  | - | - | 30.40 | 28.27 | - | - | 31.03 | 28.88 |
| NC | - | - | - |  | - | - | - |  |
|  | - | - | - |  | - | - | - |  |
|  | - | - | - |  | - | - | - |  |

**Supplementary Table 14.** Ct values from real-time PCR for analytical limit of detection of MPXV Clade II synthetic standard spiked in clinical negative matrix. NC denotes negative control.

| **Replicate** | **Batch 1** | | | | **Batch 2** | | | |
| --- | --- | --- | --- | --- | --- | --- | --- | --- |
|  | **Tube A** | | **Tube B** | | **Tube A** | | **Tube B** | |
|  | **FAM**  **B15L+F3L** | **HEX**  **B1R** | **FAM**  **SPC** | **HEX**  **ACTB** | **FAM**  **B15L+F3L** | **HEX**  **B1R** | **FAM**  **SPC** | **HEX**  **ACTB** |
| 1 | 34.39 | - | 31.35 | 28.19 | 34.96 | - | 31.54 | 28.28 |
| 2 | 34.42 | - | 31.39 | 27.94 | 34.98 | - | 31.28 | 28.28 |
| 3 | 34.29 | - | 31.33 | 27.99 | 36.13 | - | 31.43 | 28.50 |
| 4 | 34.07 | - | 31.77 | 28.13 | 35.44 | - | 31.43 | 28.46 |
| 5 | 34.33 | - | 31.41 | 27.98 | 37.14 | - | 31.63 | 28.33 |
| 6 | 34.29 | - | 31.45 | 27.99 | 35.45 | - | 31.42 | 28.31 |
| 7 | 34.23 | - | 31.49 | 28.16 | 35.56 | - | 31.35 | 28.18 |
| 8 | 34.26 | - | 31.54 | 27.90 | 35.37 | - | 31.44 | 28.27 |
| 9 | 34.37 | - | 31.26 | 27.70 | 35.39 | - | 31.45 | 27.94 |
| 10 | 35.54 | - | 31.35 | 27.99 | 35.78 | - | 31.39 | 28.22 |
| 11 | 33.58 | - | 31.36 | 28.16 | 35.82 | - | 31.41 | 27.15 |
| 12 | 34.67 | - | 31.30 | 27.88 | 35.66 | - | 31.34 | 28.06 |
| 13 | 34.07 | - | 31.45 | 28.14 | 35.85 | - | 32.28 | 27.46 |
| 14 | 34.46 | - | 31.57 | 28.10 | 35.32 | - | 31.53 | 28.20 |
| 15 | 34.67 | - | 31.34 | 27.99 | 35.69 | - | 31.43 | 28.12 |
| 16 | 34.11 | - | 31.31 | 27.84 | 31.45 | - | 31.65 | 28.20 |
| 17 | 34.38 | - | 31.51 | 28.00 | 35.72 | - | 31.48 | 28.24 |
| 18 | 34.57 | - | 31.47 | 28.21 | 39.57 | - | 32.13 | 29.13 |
| 19 | 34.45 | - | 31.42 | 28.21 | 36.54 | - | 32.06 | 29.01 |
| 20 | 34.23 | - | 31.25 | 28.12 | 35.28 | - | 31.82 | 28.68 |

**Supplementary Figure 1.** Detection probability against number of copies per reaction for MPXV Clade I (B15L+F3L and B1R assays) and MPXV Clade II (B15L+F3L assay). Curves represent fitted probit regression models. The horizontal dashed line indicates 95% detection probability. Vertical dotted lines indicate regression-estimated LoD_95_.

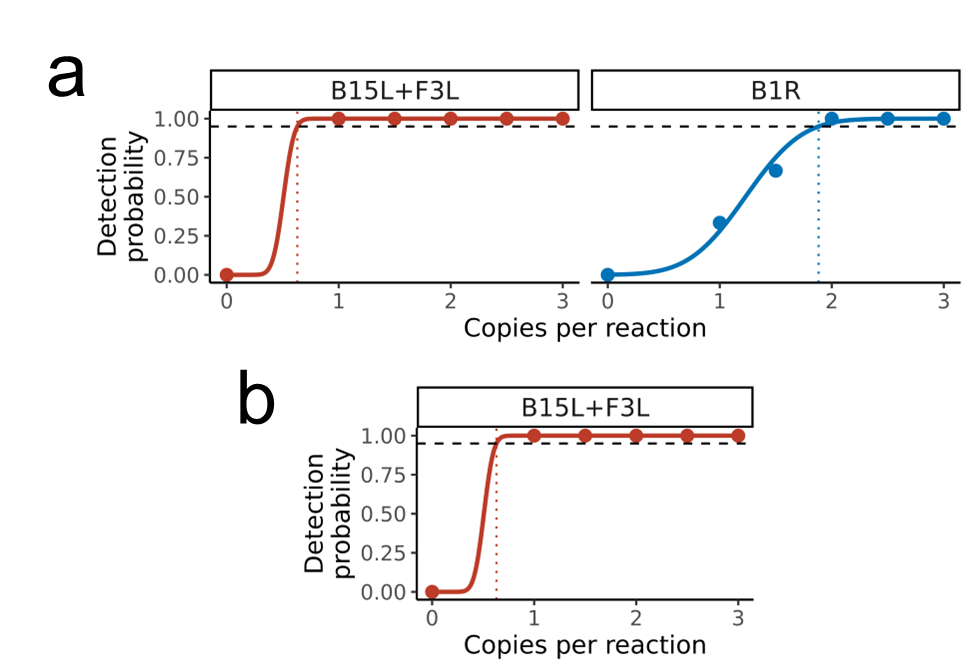

**Supplementary Table 15.** Ct values from real-time PCR for tentative limit of detection of MPXV Clade II irradiated virus spiked in clinical negative matrix. NC denotes negative control.

| **Concentration**  **(TCID_50_/ml)** | **Tube A** | | **Tube B** | |
| --- | --- | --- | --- | --- |
|  | **FAM**  **B15L+F3L** | **HEX**  **B1R** | **FAM**  **SPC** | **HEX**  **ACTB** |
| 1000 | 29.191 | - | 31.598 | 27.909 |
|  | 30.676 | - | 30.676 | 27.266 |
|  | 30.485 | - | 31.109 | 27.410 |
| 500 | 34.446 | - | 31.172 | 26.635 |
|  | 34.083 | - | 31.353 | 28.211 |
|  | 34.374 | - | 31.333 | 28.310 |
| 250 | 35.233 | - | 31.626 | 29.435 |
|  | 37.035 | - | 31.125 | 28.750 |
|  | 35.950 | - | 30.974 | 28.677 |
| 125 | - | - | 31.754 | 29.042 |
|  | - | - | 32.046 | 29.526 |
|  | - | - | 31.568 | 28.379 |
| 62.5 | - | - | 31.574 | 29.941 |
|  | - | - | 31.084 | 27.799 |
|  | - | - | 31.179 | 27.740 |
| 31.25 | - | - | 32.051 | 28.532 |
|  | - | - | 30.983 | 28.376 |
|  | - | - | 32.443 | 28.677 |
| NC | - | - | - | - |
|  | - | - | - | - |
|  | - | - | - | - |

**Supplementary Table 16.** Ct values from real-time PCR for analytical limit of detection of MPXV Clade II irradiated virus spiked in clinical negative matrix. NC denotes negative control.

| **Replicate** | **Tube A** | | **Tube B** | |
| --- | --- | --- | --- | --- |
|  | **FAM**  **B15L+F3L** | **HEX**  **B1R** | **FAM**  **SPC** | **HEX**  **ACTB** |
| 1 | 36.642 | - | 32.051 | 29.026 |
| 2 | 35.978 | - | 31.902 | 29.188 |
| 3 | 35.506 | - | 31.172 | 29.553 |
| 4 | 35.804 | - | 31.653 | 29.321 |
| 5 | 34.560 | - | 31.862 | 29.068 |
| 6 | 37.199 | - | 31.715 | 28.150 |
| 7 | 35.919 | - | 31.198 | 29.275 |
| 8 | 36.383 | - | 31.606 | 29.715 |
| 9 | 36.304 | - | 31.672 | 28.626 |
| 10 | 38.096 | - | 31.102 | 28.325 |
| 11 | 36.135 | - | 31.830 | 28.539 |
| 12 | 35.236 | - | 31.774 | 28.981 |
| 13 | 36.569 | - | 31.137 | 28.887 |
| 14 | - | - | 30.931 | 29.128 |
| 15 | 35.938 | - | 31.719 | 29.612 |
| 16 | 36.443 | - | 30.538 | 28.348 |
| 17 | 36.712 | - | 31.455 | 28.599 |
| 18 | 36.413 | - | 31.410 | 28.693 |
| 19 | 35.432 | - | 30.147 | 27.012 |
| 20 | 34.916 | - | 30.502 | 28.278 |
